# Twitter activity about treatments during the COVID-19 pandemic: case studies of remdesivir, hydroxychloroquine, and convalescent plasma

**DOI:** 10.1101/2020.06.18.20134668

**Authors:** Tymor Hamamsy, Richard Bonneau

**Author notes:** Corresponding author: Name: Tymor Hamamsy.

## Abstract

Since the COVID-19 pandemic started, the public has been eager for news about promising treatments, and social media has played a large role in information dissemination. In this paper, our objectives are to characterize the public discussion of treatments on Twitter, and demonstrate the utility of these discussions for public health surveillance. We pulled tweets related to three promising COVID-19 treatments (hydroxychloroquine, remdesivir and convalescent plasma), between the dates of February 28th and May 22nd using the Twitter public API. We characterize treatment tweet trends over this time period. Most major tweet/retweet/sentiment trends correlated to public announcement made by the white house and/or to new clinical trial evidence about treatments. Most of the websites people shared in treatment-related tweets were non-scientific, ideological media sources that leaned conservative. Hydroxychloroquine was the most discussed treatment on Twitter. There is a gap between the public attention/discussion around COVID-19 treatments and their evidence. Twitter data can and should be used for public health surveillance during this pandemic, as it is informative for monitoring adverse drug reactions, especially as many people avoid going to hospitals/doctors.

## Introduction

Since the COVID-19 pandemic started, the public has been eager for news about promising treatments. In mid-March, treatments became a focus of the White House Coronavirus briefings, with hydroxychloroquine taking center stage. In reaction to these communications, people panic-bought hydroxychloroquine, and in March over 40,000 health care professionals became first-time prescribers of the drug, with the prescribing rate in New York increasing over 40 times.[1] In March, the success of most treatments was speculative, but as of May 5th, there were 1209 COVID-19 clinical trials underway. Social media discussions about treatments during a pandemic are relatively new territory,[2-3] and with new clinical evidence constantly coming out, scientific information gets amplified to the public. While Twitter trends during the COVID-19 pandemic have been well characterized by several studies,[4-6] the extent of social media activity related to COVID-19 treatments has not been well described. In this study, we characterize trends on Twitter (San Francisco, CA), that reflect discussion of COVID-19 treatments, we show how they temporally relate to White House communications, new evidence about treatments, and also illustrate their utility for public health surveillance.

## Methods

We used Twitter’s public API, and the R package ‘rtweet’ to pull tweets mentioning several promising treatments with clinical trials underway for COVID-19, focusing on hydroxychloroquine, remdesivir and convalescent plasma.[7] We retrospectively analyzed posts made between February 28th and May 22nd, 2020, on Twitter. We calculated the number of tweets and retweets made per day mentioning each treatment. We performed sentiment analysis on these tweets using the AFFIN lexicon, and calculated the average sentiment per day for each treatment. We analyzed the urls mentioned in these tweets, counting how often different websites were shared over this time period. We searched these urls for mentions of journals/preprints, and calculated the number of retweets for each treatment that linked to research articles. We screened all hydroxychloroquine tweets for mentions of adverse drug reactions (ADRs), using the MEDRA ontology for low-level ADRs, and the spaCy NLP library for term matching. We visualized all of these data alongside major treatment/clinical trial/government announcements.

## Results

Among 1209 clinical trials, 133 interventions included hydroxychloroquine, 45 convalescent plasma, and 13 remdesivir. Between February 28th and May 22nd, we found 441,052 tweets containing “hydroxychloroquine” or “hcq”, 60,050 containing “remdesivir”, and 63,958 containing “convalescent” or “plasma” along with a treatment-related phrase.

The three hydroxychloroquine peaks appeared after Trump promoted hydroxychloroquine on March 21^st^, April 5^th^, and May 18^th^, and 31% of “hydroxychloroquine” tweets mentioned Trump. Our analysis of the links included in tweets mentioning hydroxychloroquine, showed that the top sites were mostly conservative media websites. We found 72 tweets made by congress members about treatments, and their sentiments about hydroxychloroquine were divided along party lines: average sentiment was negative for Democrats (-.41) and positive for Republicans (.88). We found 68,934 mentions of ADRs in treatment tweets, including 61,144 in hydroxychloroquine tweets (covering 10.4% of hydroxychloroquine tweets).

## Discussion

Health misinformation during this pandemic about treatments, vaccines, and prevention strategies, have been described as an infodemic.[8] Social media is playing a crucial role in information dissemination, but it has been a double-edged sword. As Figure 2 illustrates, most information about treatments reaching the public on social media comes from ideological sources, and frequently these sources spin scientific results. As an example, before evidence of the efficacy of hydroxychloroquine for treating COVID-19 came out, the white house advertised it, doctors over prescribed it, and governments stockpiled it. We’ve seen from the rapid pace of preprint articles, and premature publishing of the Surgisphere papers,[9] that science during a pandemic is messy. As evidence about treatments for COVID-19 comes out, scientific communication of the evidence, not extrapolation or speculation, will be essential.

**Figure 1.**
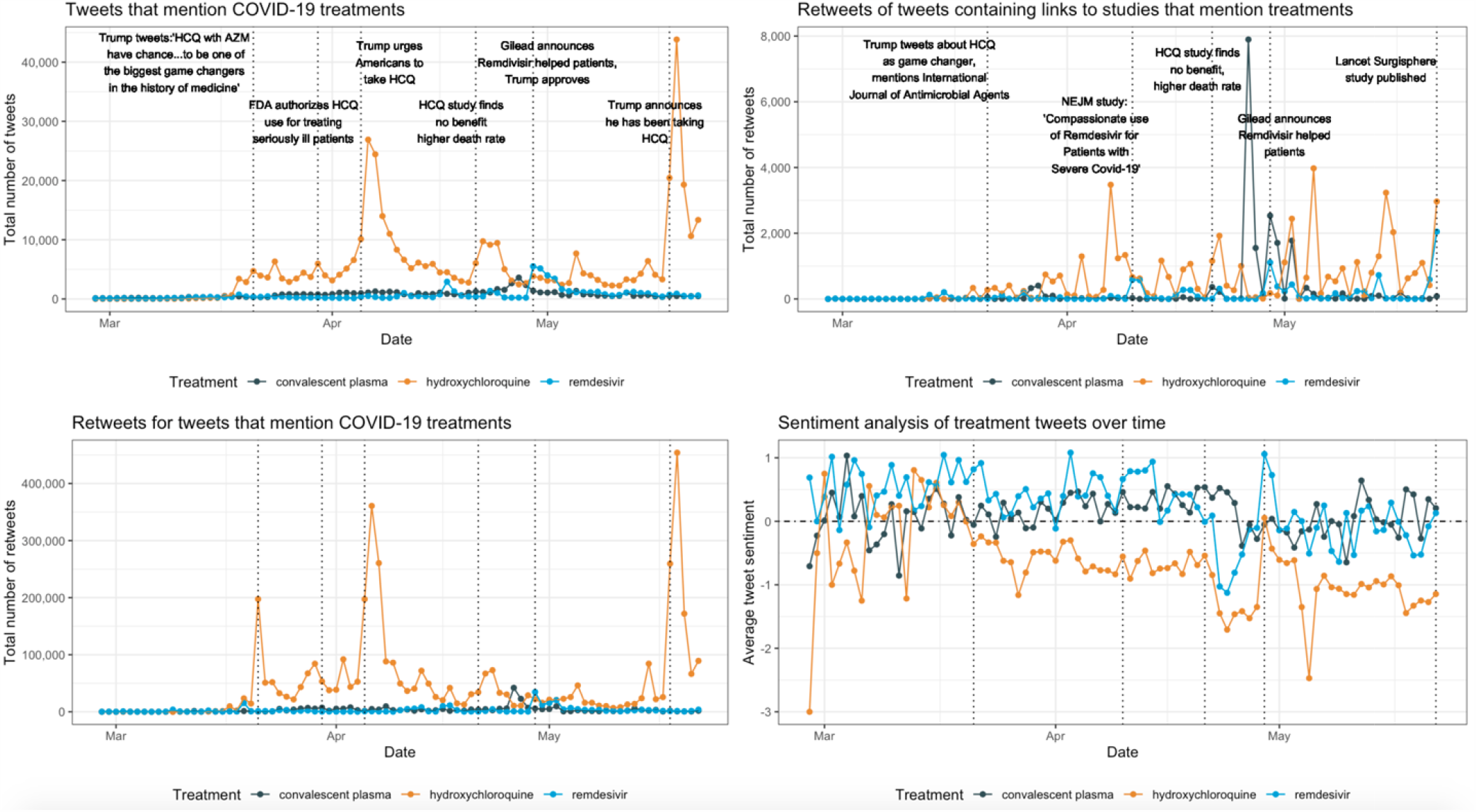
Treatment-related tweets, retweets, and sentiment trends during the COVID-19 pandemic. Tracking twitter activity for remdesivir, hydroxychloroquine, and convalescent plasma over time. After the first medrxiv study came out on April 21st showing an increased risk of death associated with hydroxychloroquine, the White House temporarily stopped promoting the drug, and hydroxychloroquine twitter activity declined precipitously, until Trump announced he had been taking it on May 18^th^.

**Figure 2:**
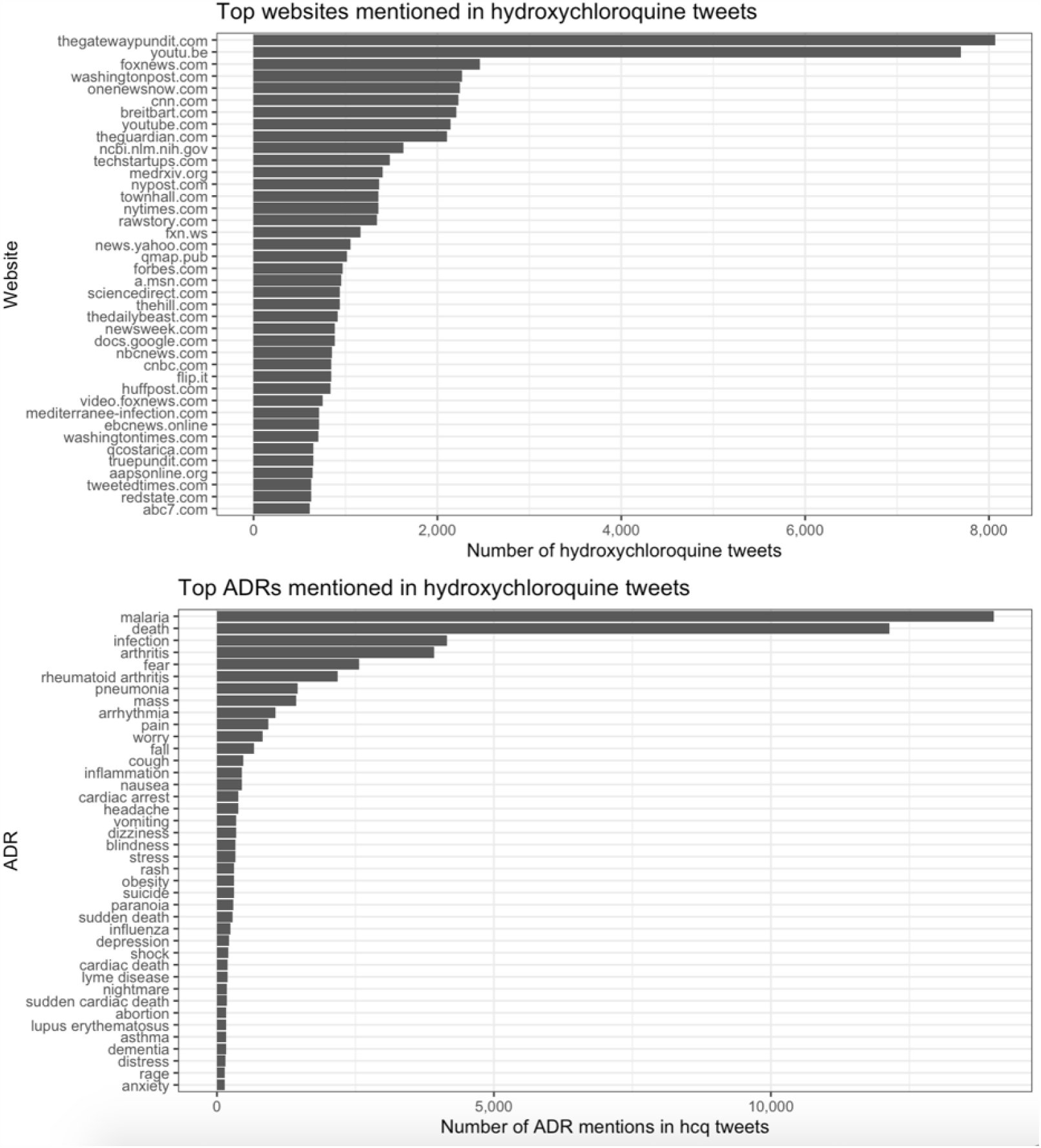
The top 40 urls and ADRs people mentioned in COVID-19 hydroxychloroquine - related tweets between February 28th and May 22^nd^. For hydroxychloroquine, many of the top websites are right-wing websites, including “thegatewaypundit”, “foxnews”, and “breitbart”. There were 620 different ADRs mentioned in hydroxychloroquine -related tweets. Given the mass promotion of this drug, and a public that is fearful of visiting hospitals during a pandemic, social media mentions of ADRs can inform public health surveillance.

## Data Availability

Data will be shared upon request. Due to the terms and conditions of Twitter, we can only share the tweet ids of tweets, and not the tweet text.

## Ethics statement

No ethical approval was needed for this cohort study that uses publicly accessible Twitter data.

